# Vaccine-induced correlate of protection against fatal COVID-19 in the old and frail during waves of neutralization resistant variants of concern

**DOI:** 10.1101/2023.02.16.23286009

**Authors:** Linnea Vikström, Peter Fjällström, Yong-Dae Gwon, Daniel J. Sheward, Julia Wigren-Byström, Magnus Evander, Oscar Bladh, Michael Widerström, Christian Molnar, Gunlög Rasmussen, Louise Bennet, Mikael Åberg, Jonas Björk, Staffan Tevell, Charlotte Thålin, Kim Blom, Jonas Klingström, Benjamin Murrell, Clas Ahlm, Johan Normark, Anders F Johansson, Mattias NE Forsell

**Affiliations:** Department of Clinical Microbiology, Umeå University, Sweden; Department of Microbiology, Tumor and Cell Biology, Karolinska Institutet, Sweden; Department of Clinical Sciences, Karolinska Institutet Danderyd Hospital, Stockholm, Sweden; Familjeläkarna Stockholm, Sweden; School of Medical Sciences, Örebro University; Clinical Studies Sweden, Forum South, Skåne University Hospital and Department of Clinical Sciences, Lund University, Sweden; Department of Medical Sciences, Clinical Chemistry and SciLifeLab, Uppsala University, Uppsala, Sweden; Division of Occupational and Environmental Medicine, Lund University, Lund, Sweden; Department of Infectious Diseases, Karlstad Hospital and Centre for Clinical Research and Education, Region Värmland and Faculty of Medicine and Health, Örebro University, Örebro, Sweden; Swedish Public Health Agency, Stockholm, Sweden; Department of Biomedical Clinical Sciences, Linköpings University, Sweden

## Abstract

**Background:** To inform future preventive measures including repeated vaccinations, we have searched for a clinically useful immune correlate of protection against fatal Covid-19 among nursing homes residents.

**Methods:** We performed repeated capillary blood sampling with analysis of S-binding IgG in an open cohort study with inclusion of nursing home residents in Sweden. We analyzed immunological and registry data collected from September 2021 with end of follow-up 31 August 2022. The study period included implementation of the 3rd and 4th mRNA monovalent vaccine doses and Omicron virus waves.

**Findings:** A total of 3012 nursing home residents with median age 86 were enrolled. The 3rd mRNA dose elicited a 99-fold relative increase of S-binding IgG among 2606 blood-sampled individuals and corresponding increase of neutralizing antibodies. The 4th mRNA vaccine dose boosted the levels 3.8-fold. Half-life of S-binding IgG was 72 days. A total 528 residents acquired their first SARS-CoV-2 infection after the 3rd or the 4th vaccine dose and the 30-day mortality was 9.1%. We found no indication that levels of vaccine-induced antibodies protected against infection with Omicron VOCs. In contrast, the risk of death was inversely correlated to levels of S-directed IgG below the 20^th^ percentile. The risk plateaued at population average above lower 35th percentile of S-binding IgG.

**Interpretation:** In the absence of neutralizing antibodies that protection from infection, quantification of S-binding IgG post vaccination may be useful to identify the most vulnerable for fatal Covid-19 among the oldest and frailest. This information is of importance for future strategies to protect vulnerable populations against neutralization resistant variants of concern.

**Funding:** Swedish Research Council, SciLife, Knut and Alice Wallenberg Foundation and Vinnova.

## INTRODUCTION

Prior to the rollout of SARS-CoV-2 vaccines, Covid-19 caused substantial morbidity and mortality worldwide, particularly in older individuals. In Sweden, elderly care services are provided in nursing homes staffed and equipped to maintain good quality of life ^1-3^. Approximately 80% of the individuals in this population are over the age of 80 and many have comorbidities including dementia^4,5^. Early in the pandemic, diagnosed SARS-CoV-2 infection was associated with a 40% excess 30-day mortality among residents at Swedish nursing homes^5^. Accordingly, nursing home residents were prioritized for vaccination against Covid-19, and 88% had received two vaccine doses as of 1 April 2021^6^. A majority of these individuals received two doses of BNT162b2 (Pfizer/BioNTech) as their primary vaccination regimen.

A two-dose SARS-CoV-2 mRNA vaccine regimen induces SARS-CoV-2 Spike-directed antibody-responses in elderly individuals ^7-11^, though advanced age has been associated with weaker and less durable responses ^8,12,13^. Low titer antibody responses may be even more vulnerable to antibody escape by emerging variants of concern^14^, raising concerns about the longevity of protection against infection and disease among older nursing home residents. To counteract the waning of specific immune responses, repeated vaccinations have been administered at Swedish nursing homes. A third dose was administered to residents at nursing homes approximately eight months after the primary two-dose regimen. A fourth dose was then recommended to individuals >80-year-old and to all nursing home residents approximately five months after the third dose. For both the third and fourth doses, the vaccine regimens comprised of either BNT162b2 or a half-dose of mRNA-1273 (Moderna). The beneficial effect of these booster doses in reducing infections and mortality in the general population in Sweden has previously been demonstrated^15^.

A measurable correlate of protection against Covid-19 mortality would be extremely valuable to identify individuals and cohorts at risk for severe or fatal outcome after a SARS-CoV-2 infection. The aim of the study was to evaluate if levels of vaccine-induced circulating IgG could predict susceptibility for infection or future fatal Covid-19 from infection with variants of concern among nursing home residents. Thus, in a cohort of nursing home residents, we characterized longitudinal Spike (S) -binding antibody responses with high resolution after three or four vaccine-doses and then correlated the results with registry-based data on infections and mortality on an individual basis.

## RESULTS

### Population and setting

A total of 3012 nursing home residents from 114 nursing homes were enrolled in an observational open cohort study in Sweden. The inclusion period was from 16 September 2021 to 13 April 2022, with inclusion start before the roll-out of a third Covid-19 vaccine dose with a mRNA vaccine. Characteristics of the population at baseline, vaccinations, and capillary sampling for repeated analyses of SARS-CoV-2 antibodies in blood are described in Table 1. Both metropolitan and less densely populated regions were represented (Figure 1A), and the residents comprised a higher proportion of women than men above 75 years (Figure 1B). A high weekly incidence within the cohort of PCR-verified SARS-CoV-2 infections with Omicron variants of concern (VOC) facilitated analyses of outcomes with relation to immune responses (Figure 1C). After the laboratory analyses, at least one S-binding IgG observation in blood was available for 2607 study subjects and there were a total number of 6958 S-binding IgG observations within the cohort.

**Table 1.** Characteristics of the study population.

| Characteristic | All regions<br>(N=3012) | Northern and<br>central regions<br>(N=2174) | Metropolitan<br>regions<br>(N=838) |
| --- | --- | --- | --- |
| Median age (range) -yr | 86 (41 - 105) | 86 (41 - 105) | 85 (59 - 103) |
| Female sex -no. (%) | 1865 (64.1) | 1374 (63.2) | 547 (65.3) |
| History of PCR-verified SARS-CoV-2 at inclusion -no. (%) | 405 (13.4) | 196 (9.0) | 209 (24.9) |
| History of vaccination at inclusion -no. (%) * |  |  |  |
| 1 dose | 39 (1.3) | 28 (1.3) | 11 (1.3) |
| 2 doses | 1829 (60.7) | 1756 (80.8) | 73 (8.7) |
| 3 doses | 1144 (38.0) | 390 (17.9) | 754 (90.0) |
| Interval between vaccine doses -days * |  |  |  |
| Dose 1 - 2 median (range) | 28 (14 - 477) | 28 (14 - 477) | 21 (17 - 416) |
| Dose 2 - 3 median (range) | 245 (57 - 555) | 242 (100 - 555) | 252 (57 - 532) |
| Dose 3 - 4 median (range) | 146 (69 - 353) | 143 (69 - 350) | 149 (82 - 353) |
| Median inclusion date | 08 Oct 2021 | 05 Oct 2021 | 22 Nov 2021 |
| Subjects entering/leaving study -no. |  |  |  |
| 16 Sept - 27 Nov 2021 | 2563 / 160 | 2047 / 146 | 516 / 14 |
| 28 Nov 2021 - 27 Feb 2022 | 193 / 265 | 71 / 201 | 122 / 64 |
| 28 Feb - 13 April 2022 | 256 / 103 | 56 / 81 | 200 / 22 |
| Subjects donating blood sample -no. |  |  |  |
| Sampling period 1: 28 Sept - 27 Nov 2021 | 1570 | 1570 | 0 |
| Sampling period 2: 28 Nov 2021 - 19 Jan 2022 | 1950 | 1474 | 476 |
| Sampling period 3: 28 Feb - 05 May 2022 | 1791 | 1258 | 533 |
| Sampling period 4: 19 May - 23 July 2022 | 1647 | 1218 | 429 |
| No. of blood samples donated per person -no. |  |  |  |
| One | 496 | 331 | 165 |
| Two | 654 | 460 | 194 |
| Three | 798 | 503 | 295 |
| Four | 690 | 690 | 0 |

**Figure 1.**
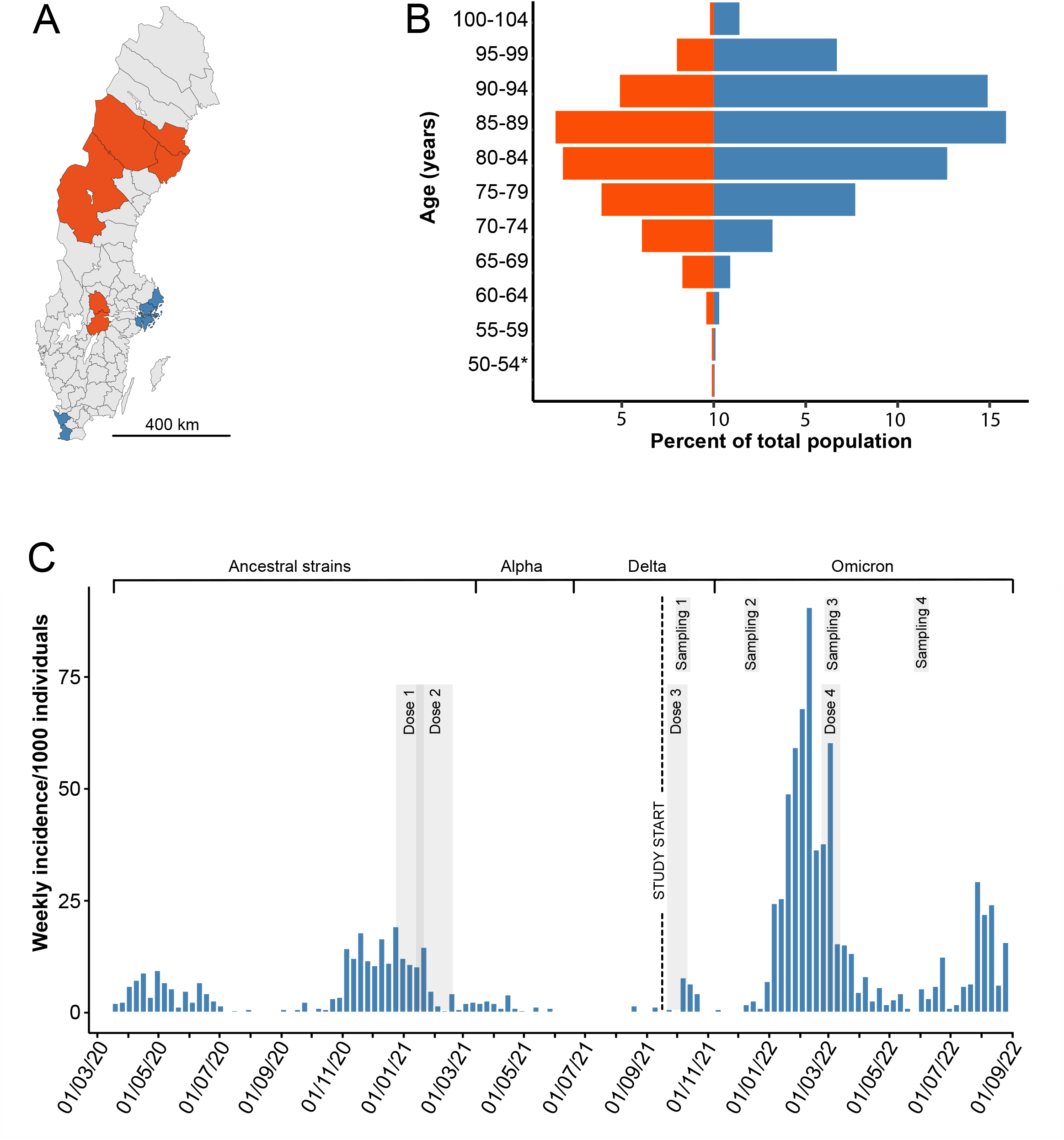
Location of nursing homes, age and gender characteristics, and the progression of PCR-verified SARS-CoV-2 infections in the study population of 3012 study subjects. Panel A shows locations in Sweden of nursing homes with recruitment of study subjects in two metropolitan areas (Stockholm and Malmö; blue color) and three less densely populated regions (Västerbotten, Jämtland-Härjedalen and Örebro; red color). Panel B shows proportions of the total population stratified per age-group of women (blue bars) and men (red bars) An asterisk indicates that one 41-year-old study subject was included. Panel C shows PCR-verified SARS-CoV-2 incidence among the study subjects over the entire pandemic. Timing of vaccine doses, study start, blood sampling periods, dominating virus variant, periods of vaccination, and blood sampling are indicated. For vaccination and sampling, a gray-shaded interval represents that 80% of the population was completed.

### Robust induction of S-directed IgG after a third and fourth dose of mRNA vaccination among nursing home residents

The vaccine coverage was 95.9% for the third dose and 80.1% for the 4fourth dose. We found that >90% of the vaccinees received the third dose within 41 calendar days (median date October 13, 2021), and >90% received the forth dose within 37 calendar days (median date March 7, 2022). We analyzed S-binding IgG levels before and after the third mRNA vaccination dose with either the standard dose of BNT162b2 or a half-dose mRNA-1273 given after a primary 2-dose regimen of vaccination with BNT162b2 eight months before (Figure 2A). Previously infected individuals had higher anti-S levels both before and after vaccination. Infection-naïve individuals displayed a 99-fold relative increase in anti-S IgG levels (median AUC 352 at day-40 to -1 before dose 3 (N=653); median AUC 34,978 at day 14 to day 28 after dose 3 (N=185)). By logarithmic linear regression, we estimated a mean anti-S IgG level decline between day 20 and 80 post dose 3 at 1.3% per calendar day, equivalent to a half-life of 70.2 days. The increase in S-binding IgG after dose 3 was reflected in an increased capacity of serum antibodies to outcompete ACE-2 binding to the Wu-1 S-protein *in vitro* (Supplemental Figure 1A).

**Figure 2.**
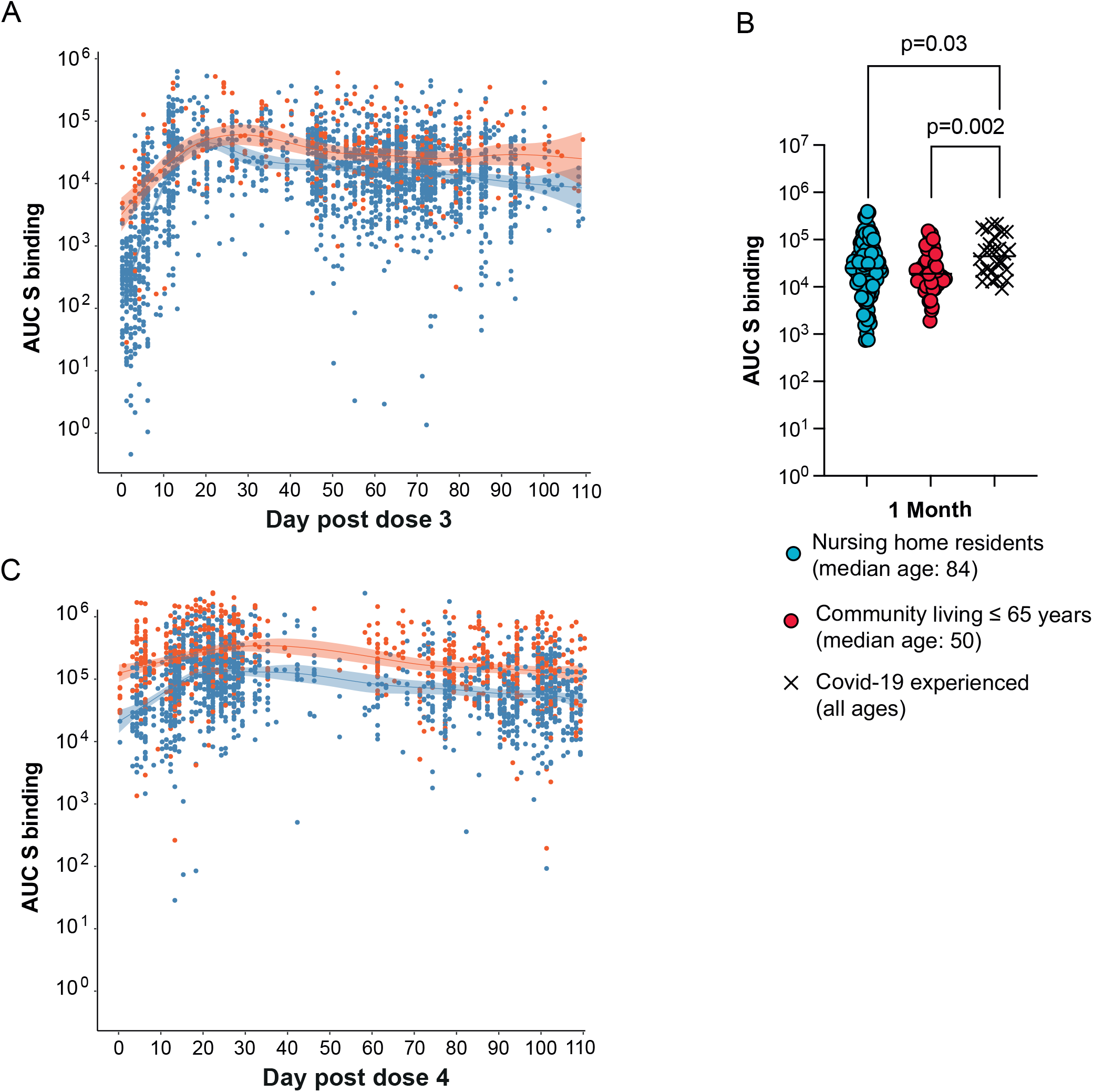
Dynamics and projection of S-directed IgG responses. Panel A shows S-directed IgG levels with respect to the 3^rd^ dose mRNA-vaccination, as assessed by repeated capillary blood sampling of previously uninfected (blue) or previously infected (red) nursing home residents. Day 0 is the day of vaccination. Shading indicates 95% confidence intervals. Panel B shows comparison of S-directed IgG in the nursing home population (blue) and community living individuals up to the age of 65 (red) and those previously infected with SARS-CoV-2 covid-19 from both groups (black crosses). Median for each group is shown. Panel C shows S-directed IgG levels before and after the 4^th^ dose mRNA-vaccination of the nursing home population. Each dot represents one study subject.

By investigating capillary blood samples taken approximately 1 month after the third dose, we found no difference in S-binding levels between previously uninfected nursing home residents and previously uninfected community living individuals up to the age of 65. (Figure 2B, supplemental table 1). For both groups, the levels of S-binding IgG were significantly lower when compared to individuals with a confirmed SARS-CoV-2 infection prior to the third dose.

In comparison with the third dose, the fourth dose led to a more modest 3.8-fold relative increase in circulating S-binding IgG (median AUC 14,587 from day -40 to -1 before dose 4 (N=364); median AUC = 55,729 from day 14 to 28 post dose 4 (N=701)) (Figure 2C), again reflected by an increased capacity of serum antibodies to outcompete ACE-2 binding to the Wu-1 S-protein (Supplemental Figure 1B). There was no significant difference in the decline of S-binding IgG after 4th vaccine dose as compared with after the third vaccine dose (p-value 0.579).

By generalized additive modelling, we found that median AUC of S-binding IgG after the third mRNA vaccine dose was 1.75 times higher with a half dose mRNA-1273 (N=202) than with BNT162b2 (N=1576) (p-value <0.0001) (Supplemental figure 2). The modelling revealed no significant gender difference in the response levels (p=0.525). Given the lack of a formal randomization between the vaccine brands in the study design and that our goal was to study overall vaccine responses in the nursing home population, we did not deem that further stratification of data based on vaccine-moiety given was warranted.

### Comparative analysis of vaccine-induced S-specific antibodies after a 4^th^ dose between old and young

Venous blood sampling allowed us to do a more detailed serum analysis on days 0, 7-10 and 30 after the fourth vaccine dose. We focused on SARS-CoV-2 unexposed individuals and found that the fourth dose had induced similar increases in S-binding IgG in serum from old persons at nursing homes (median AUC_pre4_ 12398 to AUC_post4_ 66359), community living persons aged >66 (median AUC_pre4_ 20885 to AUC_post4_ 77142) or community living persons up to the age of 65 (median AUC_pre4_ 10843 to _AUCpost4_ 67741) (Figure 3A, supplemental table 2). This was reflected by significantly increased *in vitro* neutralizing titers against the original SARS-CoV-2 strain among old persons at nursing homes, community living persons over the age of 65 or up to the age of 65 (5.3x, 3.7x or 6.2x increase, respectively) (Figure 3B). Statistical analysis demonstrated that samples from 5 individuals under the age of 65 had slightly higher median neutralization titer than samples from 16 nursing home residents after a fourth dose, whereas 27 community-living old individuals had higher neutralization titers than the nursing home residents prior to the fourth dose. We could also demonstrate that the increase in homologous neutralization was associated with enhanced neutralization of Omicron (BA.2, BA.2.75 and BA.5) by serum from nursing home residents (Figure 3C).

**Figure 3.**
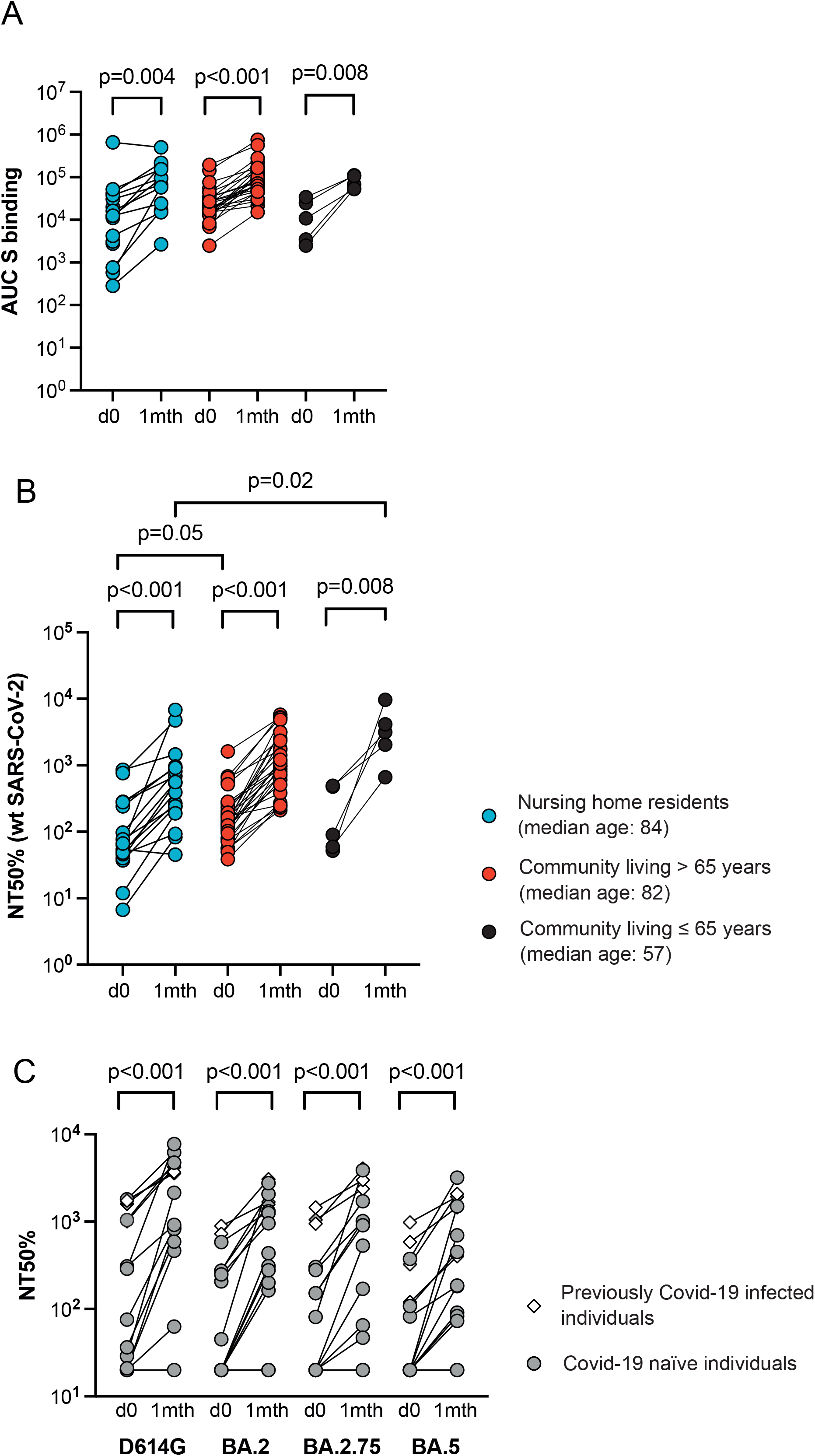
Detailed effect of booster vaccination. Samples from nursing home residents (blue circles), community living older individuals (red circles) or community living younger individuals (≤65 years, black circles) were assessed for S-binding IgG (Panel A) or authentic neutralization against Wu-1 isolate of SARS-CoV-2 (Panel B). S-binding is shown as the area under the curve for each sample (AUC) and the reciprocal serum dilution at which 50% neutralization was achieved (NT50%). Panel C shows neutralization of SARS-Cov-2 S-pseudotyped virus by serum from nursing home residents on day 0 (open circles) and 30 post (black circles) dose 4. Reciprocal serum dilution where 50% neutralization of SARS-CoV-D614G, BA.2, BA.2.75 or BA.5 is shown (NT50%). Each dot represents one individual.

### Infection with relation to booster doses at nursing homes

After study start 16 September 2021, 25% of included study subjects (765/3012) acquired a primary PCR verified SARS-CoV-2-infection. Twenty-seven study subjects acquired a second PCR verified infection. None was registered with three or more PCR-verified infections during the pandemic. Thirty-two subjects acquired the primary PCR verified infection during the delta virus dominance from 16 September 2021 to 31 December 2021 and 733 acquired the primary infection during the remainder of the study, where Omicron variants BA.1 and BA.2 were dominant until superseded by BA.5 in June 2022.

We determined if vaccine-induced antibody levels had an impact on susceptibility to BA.1 or BA.2 infection by comparing projected antibody levels among study subjects that acquired PCR-verified infection with the levels among study subjects not acquiring infection. Among 528 previously infection-naïve study subjects vaccinated with three mRNA doses, we found no difference in levels of S-specific IgG to study subjects that subsequently acquired infection over the corresponding 14-day periods after the third mRNA vaccine dose (Supplemental figure 3).

### Antibody levels in relation to mortality at nursing homes

Throughout the study period, 781 study subjects died: 243 from 16 September to 31 December 2021, and 538 from 1 January to 31 August 2022. A main reason for that some study subjects did not donate a blood sample after inclusion was short life expectancy: median time to death for 2607 subjects donating or 405 subjects not donating blood were 149 and 37.5 days after inclusion, respectively. From study start until the end of register follow-up 30 September 2022, 65 deaths occurred within 30 days after a PCR-verified SARS-CoV-2 infection, five during Delta and 60 during the Omicron waves.

Among the 528 individuals that acquired their first SARS-CoV-2 infection after the third or the fourth vaccine dose and where we had an appropriately timed analysis of S-directed IgG available, the 30-day mortality was 9.1% (48 deaths/528 cases). This compared with a non-covid-related 30-day mortality of 2.6% (27/1056) among control individuals sampled at the same day (Figure 4A). We found no difference in 30-day mortality between individuals that acquired infection after the third (32 deaths/362 cases) or the fourth mRNA vaccine dose (16 deaths/166 cases) (Kaplan-Meier logrank test, p=0.75) (Supplemental figure 4)

**Figure 4.**
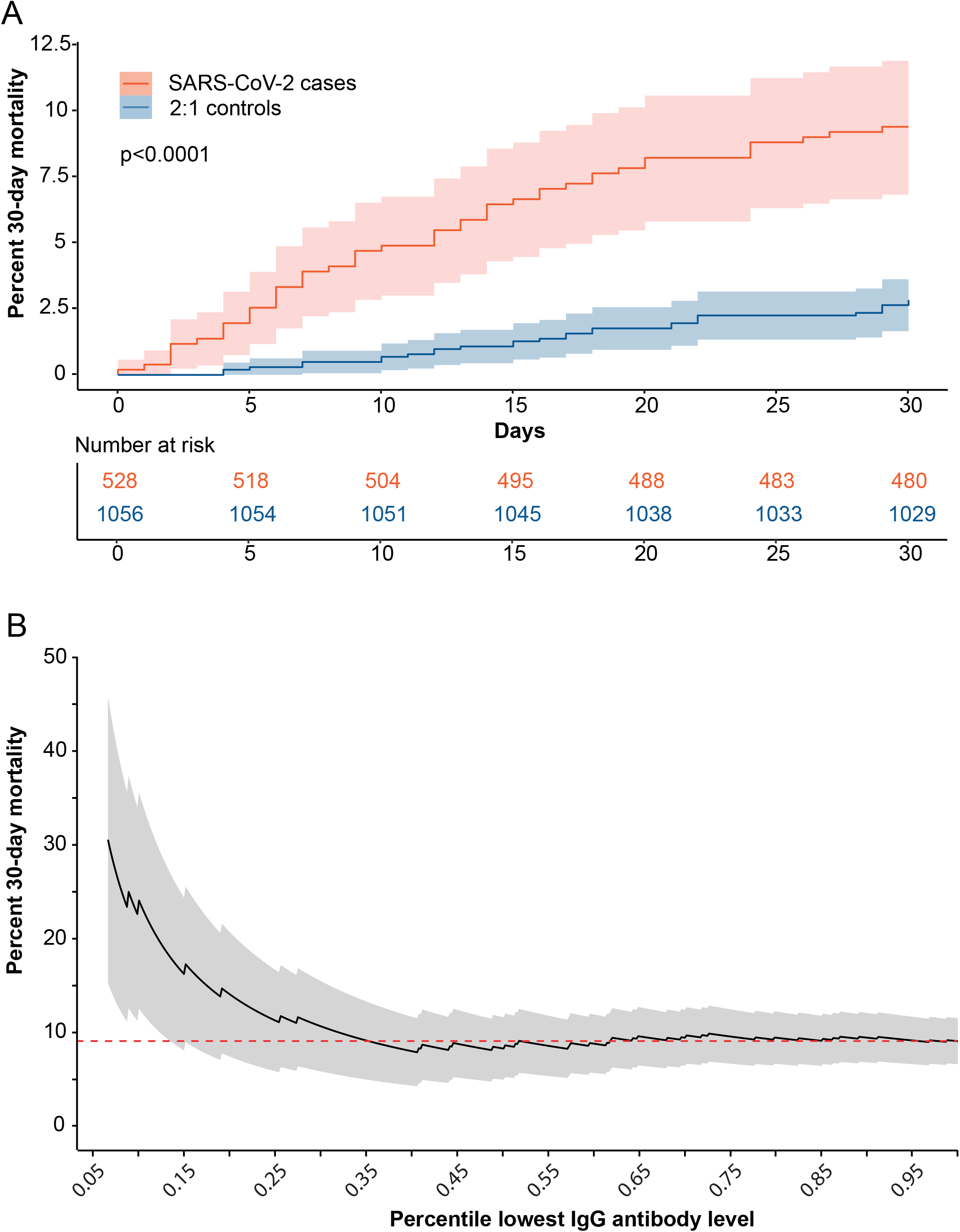
SARS-CoV-2-associated 30-day mortality. Panel A shows cumulative 30-day SARS-CoV-2-associated mortality among 528 previously infection naïve nursing home residents that were vaccinated with three or four mRNA vaccine doses before the infection (red). Control study subjects (blue) were sampled 2:1 adjusted to calendar time. Panel B shows the SARS-CoV-2 related 30-day mortality with respect to S-binding IgG levels in the nursing home population. The mortality is shown in relation to the percentile range of lowest antibody levels among the 528 SARS-CoV-2 cases and refers to antibody levels at day 60 post vaccine dose 3.

The 30-day mortality risk was higher among the 528 previously SARS-CoV-2 infection naïve individuals with low levels of S-directed IgG and this effect plateaued to reach the population average at approximately the 35th percentile (Figure 4B). Sensitivity analyses using stepwise Kaplan-Meier curves showed that an increased 30-day mortality effect was significant below to the 20^th^ percentile of circulatory S-binding IgG (Supplemental figure 5). For example, individuals with the 10th percentile lowest S-binding IgG (N=53) had projected AUCs at <479 at day 60 post the 3rd mRNA vaccine dose and 24.5% (13/53) 30-day mortality after PCR-verified SARS-CoV-2 infection. This contrasted with 7.4% (35/475) mortality among individuals with a measured AUC ≥479 (N=475). We further modelled 30-day mortality using Cox proportional hazards regression to take additional risk factors for fatal SARS-CoV-2 into account. By this analysis, we found that nursing home residents with an AUC <479 were at increased risk (hazard ratio 3.61, p-value <0.0001), as were residents infected with SARS-CoV-2 (hazard ratio 2.70, p-value <0.0001) or those of male gender (hazard ratio 2.19, p-value 0.0086). Finally, and as well established, we found that each one-year increase in age increased the risk for Covid-19-related mortality (hazard ratio 1.04, p-value 0.0216) (Supplemental table 3). We found no significant interaction effect between an AUC<479 and increased risk to acquire a SARS-CoV-2 infection (p-value 0.1219). Taken together, the Cox modelling suggests that on top of an increased mortality imposed by SARS-CoV-2 infection there is an overall increased 30-day mortality among individuals with low S-antibody responses.

## DISCUSSION

A nationally coherent vaccination scheme and a strong clinical staff network at nursing homes and a proven strategy to collect capillary blood samples^16^ allowed us to prospectively analyze vaccine-induced humoral immunity and to combine these data with high-quality register data on the individual level. The nursing home population in Sweden was continuously surveilled by rigorous PCR-based diagnostics of suspected SARS-CoV-2 infections during the study, in accordance with national and regional recommendations. This facilitated meaningful follow-up of 30-day mortality from SARS-CoV-2 infection and a comparison with vaccine-induced antibody responses.

By a detailed analysis of immune responses, we verified an increase in cross-neutralizing antibodies against Omicron lineages BA.1, BA.2.75, and BA.5 after the fourth dose of mRNA vaccination. This is consistent with increased antibody-mediated neutralization of the homologous original SARS-CoV-2 strain to increase protection also against in vitro infection with VOCs ^14,17^. The relatively lower boost effect of the 4^th^ vaccination, as compared with the 3^rd^ may be explained, at least in part, by a previously described feedback mechanism where pre-existing specific antibodies can limit re-activation of humoral immunity ^18-20^. Our analysis demonstrated that both the 3^rd^ and the 4^th^ vaccine dose induced a boost effect on SARS-CoV-2 S-binding that was similar between the study cohort and comparator groups of community living individuals. We also found that the 4^th^ vaccine dose induced significantly higher neutralizing antibody titers in sera from five younger community living individuals than in sera from nursing home residents, but not from community living individuals over the age of 65. These data support that age related variability of the immune response to two doses of mRNA vaccination had remained even after additional booster doses^8,12^

The study period included the first omicron waves in Sweden and started at a time point when the third mRNA vaccination was distributed to nursing home residents. We found no pattern supporting that higher levels of circulating S-binding IgG protected against SARS-CoV-2 infection. This may be attributable to the described significant escape from neutralizing antibodies by Omicron^21,22^. It has previously been shown that vaccine-induced neutralizing antibodies provided a correlate of protection against infection with the ancestral strain or relatively homologous VoCs^23,24^. Our results complement these data and demonstrates that S-binding IgG antibody levels provide a correlate of protection against fatality related to infections with VoCs.

Importantly, our analyses identify a threshold titer of anti-S IgG in capillary blood taken after the 3^rd^ vaccine dose below which there was significantly increased risk of fatal Covid-19. We found that the study subjects having the 20^th^ lowest percentile of S-binding IgG 60 days after the 3^rd^ vaccine dose had a significantly increased risk of fatal Covid-19. The risk effect included study subjects that in addition to the 3^rd^ vaccine dose also received the 4^th^ dose. We conclude that S-specific IgG against the original SARS-CoV-2 strain can serve as a correlate of protection against fatal Covid-19 after infection with SARS-CoV-2 variants that demonstrate significant immune escape from neutralizing antibodies. An additional interesting observation was that Covid-19-related fatalities occurred within 20 days of a confirmed SARS-CoV-2 infection in all individuals with low S-specific IgG in blood. This could indicate a role of non-neutralizing antibodies to control the infection, as previously suggested^25^. Importantly, individuals with low S-specific IgG in circulation represented a minority, and our data demonstrate that between 70-80% of all individuals at Swedish nursing homes have achieved maximal vaccine-induced protection against fatal Covid-19 after 3 or 4 doses of monovalent mRNA vaccination.

The study has certain limitations; Nursing home residents at a terminal stage of life were not included in the study, we lacked detailed information comorbidities and data to describe severity of infection was not accessible. Hence, this study was limited to an understanding of diagnosed SARS-CoV-2 infections and 30-day mortality. Moreover, we could not perform neutralization assays on the small sample volumes acquired from capillary blood, and we lacked information on mucosal Spike-specific IgA levels that may be importance to for protection against infection ^26,27^. These limitations highlight the need for further studies to address these issues and provide a more comprehensive understanding of the impact of vaccination and correlates of protection against SARS-CoV-2 variants of concern.

In conclusion, the data presented here demonstrate that levels of vaccine-induced antibodies in capillary blood can act as a correlate of protection against fatal Covid-19 among elderly at nursing homes after infection with neutralization resistant VOCs. Together with information of S-specific antibody-decline, these data may be used to predict the long term overall protective effect after vaccination and provide information for the implementation of future strategies to protect high-risk groups against VOC-related mortality.

## MATERIALS AND METHODS

### Study design and population

Study subjects living in nursing homes were included if they had received at least one Covid-19 vaccine dose and volunteered to participate in donating capillary blood for immunological analyses and linking with registry data. Informed consent was obtained via an oral and written procedure either the individual or their legal representative (usually next-of-kin) at the nursing home. We used a prospective open cohort design where study subjects could choose to be included every third month, or if already included, continue to donate a capillary blood sample. To enable comparison of immunological responses in nursing home residents, we included community living study subjects under and over the age of 65. Consent procedures, ethical permits and more details on design and population is found in Supplemental methods section.

### Data sources, and linkage

Immunological data from individuals were linked with data in the National vaccine-register (Swedish Public Health Agency), the register containing mandatory reports of all SARS-CoV-2 laboratory and clinical diagnoses in Sweden (SmiNet, Swedish Public Health Agency), and vital status in the total population register (The Swedish Tax Agency) using the person identity number given to all persons registered in Sweden.

### Sampling strategies

The capillary sampling strategy utilized has been previously described^16^, with few modifications. Briefly, individual capillary blood sampling kits were sent to study participants via postal services and sampling was conducted by finger pricking by the individual study subject or assisted by an employee at the nursing home.

### S-binding antibody responses, N-binding and ACE-competition

Circulating IgG against vaccine-derived S proteins were measured in serum or eluted capillary blood as previously described^28^. Binding antibodies to the N protein of SARS-CoV-2 and antibodies that block ACE-2 binding to the S protein of SARS-CoV-2 Wu-1 were measured using the V-PLEX SARS-CoV-2 Panel 2 (Meso Scale Diagnostics, Maryland, USA) for IgG and ACE-2 according to the manufacturer’s instruction at the SciLifeLab Affinity Proteomics Unit (Uppsala, Sweden). More details on the immunological methods are found in Supplemental methods section.

### *In vitro* neutralization assays

Assessment of the potency of serum to neutralize authentic SARS-CoV-2 *in vitro* was performed under BSL-3 conditions as previously described^28^ Pseudovirus neutralization was performed as previously described^29^ with modifications as described in Supplemental methods section.

### Outcomes

We assessed three outcomes: humoral immune responses to SARS-CoV-2 before and after a vaccine dose, SARS-CoV-2 infection as determined by a SARS-CoV-2 RT-PCR-positive clinical specimen, and SARS-CoV-2–associated mortality as defined by death occurring within 30 days after a SARS-CoV-2-infection. Outcomes were measured from 16 September 2021 to 31 August 2022 with follow-up of deaths until 30 September 2022.

### Data processing

Data processing was carried out by R version 4.1.2 (2021-11-01) – “Bird Hippie”. The code is available at https://github.com/johanssonresearch/Correlate-of-vaccine-protection-against-fatal-COVID-19

### Covariates

We adjusted analyses at all time points for previous SARS-CoV-2 infections verified by SARS-CoV-2 PCR-positivity. We further adjusted the analyses for date of birth, gender, and nursing home caregiver.

### Statistical analyses

Vaccine coverage was calculated based on the vital population at the data for the first administration of the 3^rd^ and 4^th^ mRNA vaccine dose, respectively. We used a cubic spline model with 8 and 10 knots for vaccine dose 3 and 4, respectively, to capture and plot the non-linear trend of S-directed antibody levels in blood over time. We used a generalized additive model with cubic splines to test for difference in antibody levels after dose 3 between the vaccines BNT162b2 and mRNA-1273 and for interaction between the vaccine and the decline in AUC over time with adjustment for age and gender. We used observations of S-binding IgG AUC in infection-naïve study subjects from day 20 to day 80 after dose 3 to approximate the decline by logarithmic regression. SARS-CoV-2-associated 30-day mortality with and without consideration of S-binding IgG antibody levels was analyzed by cumulative Kaplan-Meier estimates and survival at day 30. We used a case-control design with two random controls sampled the same day as each SARS-CoV-2 as described in the technical appendix 1. For analyses of S-directed antibody level as a correlate of protection against SARS-CoV-2-associated 30-day mortality, we classified the 528 Covid-19 cases into two groups: normal and low antibody responders. Cases with an S-directed antibody AUC lower than given percentile (2.5, 5, 7.5, 10, 12.5, 15, 17.5, 20) at day 60 post vaccine dose 3 were classified as low responders and the other cases as normal responders. We used a Cox proportional hazard model to estimate the risk of death within 30 days adjusting for *covid* (PCR-verified SARS-CoV-2 infection or no infection), *responder* (low or normal AUC-values), *age* (in years) and *gender* (male of female assigned at birth). We used Graphpad Prism version 9.x for statistical analysis of data in figures 2B and 3. We used non-parametric Mann-Whitney test, Wilcoxon matched-pairs signed rank test for group wise comparisons, or non-parametric Kruskal-Wallis test as appropriate. We used Dunn’s multiple comparison test for post-hoc analyses.

## Supporting information

Suppelemental materials Vikstrom Fjallstrom

## Data Availability

All data produced in the present work are contained in the manuscript

## Acknowledgements

We would like to thank the Swedish Public Health Agency for their support of this study and the Clinical Research Center at the University Hospital of Northern Sweden for assistance with sampling. We thank the coordinating study nurses Maria Grimfelt and Ida-Lisa Persson, and regional study nurses. We would also like to thank Maj Järner, Jenny Gardfjäll, Jasmin Nazemroya and Andy Dernstedt for assistance with sample processing. Finally, we would like to acknowledge all study subjects for their participation in this study, and the personnel at the nursing homes for invaluable support.

