## Supplementary material for "Vaccine-induced correlate of protection against fatal COVID-19 in the old and frail during waves of neutralization resistant variants of concern": Suppelemental materials Vikstrom Fjallstrom

**Supplementary information**

Vikström et al, Vaccine-induced correlate of protection against fatal COVID-19 after heterologous SARS-CoV- 2 infection of nursing home residents.

**Content**

Supplemental materials and methods
Supplemental table 1
Supplemental table 2
Supplemental table 3
Supplemental figure 1
Supplemental figure 2
Supplemental figure 3
Supplemental figure 4
Supplemental figure 5
Supplemental figure 6
References

### Supplemental materials and methods

#### Study design, population and ethical permits

All managers at all nursing homes in the regions Västerbotten, Jämtland-Härjedalen and Örebro were informed about the study through clinical networks set up for local healthcare governance. Nursing home managers in the metropolitan area of Stockholm and Malmö were informed through the healthcare provider Familjeläkarna AB or the research network Clinical Studies Sweden (Forum Söder), respectively. The managers agreed to help in distributing study information and allowed employed care workers to assist study subjects if needed. A legal representative of a study subject could consent to participate in the study in cases where the individual could not do this due to dementia or for other reasons. Donating capillary blood samples and being included also provided that the individual did not express concerns verbally or with body language. To enable comparison of immunological responses in nursing home residents, we also included community living study subjects under and over the age of 65 through an open cohort study of covid-19 vaccination[1]. Study subjects under the age of 65 was included via an on-going open cohort study of SARS-CoV-2 infection and vaccination of health care workers (the COMMUNITY study[1]). All study subjects were included by informed consent. The study was approved by the Swedish Ethical Review Authority (decisions 2020-01653 or 2021-00055 including amendments 2021-01397, 2021-02328,2021-03937 and 2022-00564-02).

#### Sampling strategies

Capillary blood was added to a Dried Blood Spot sample card (qDBS, Capitainer AB) that comprised two filter paper discs, each containing 10µl of blood after successful sampling. The sample cards were returned to the laboratory via postal service. Blood was eluted in PBS from the filter paper disc and stored at +4°C until analyzed for SARS-CoV-2 S-directed IgG as previously described[2]. Venous blood sampling was performed as previously described[1,3].

#### S-binding antibody responses, N-binding and ACE-competition

We used a 5-fold dilution series of serum or capillary blood to detect anti-IgG SARS-CoV-2 S-protein was Brifly, serum or eluates from capillary blood sampling cassettes were added to S-protein coated wells and incubated over-night at +4°C. After washing the wells, goat-anti human IgG alkaline phosphate conjugated antibody (#A18814, Thermo Scientific) diluted 1/8000 was added 1 hour at +37°C. After washing, 100 µl of Phosphatase Substrate 1mg/ml (#S0942, Sigma-Aldrich) was added to each well and incubated for 30 min at +37°C and then the colorimetric reaction was stopped with 50 µl 3M NaOH per well and absorbance was measured spectrophotometrically at 405 nm with microplate reader Tecan Sunrise. Specific IgG levels were assessed as the area under the curve (AUC) from the titers, with cut off OD value at 0.15 (Graphpad Prism 9.4.1). For binding N-antibody measurement, extracted qDBS samples were diluted 1:2500 and detected using a SULFO-TAG conjugated mouse monoclonal anti-human IgG antibody. For inhibition of ACE-2 spike binding, the samples were diluted 1:5 to allow competition with the recombinant human ACE2 conjugated with SULFO-TAG for binding to the spike antigen. The plates were analyzed on a MESO QuickPlex SQ 120 instrument, an electrochemiluminescence (ECL) reader measuring the light emitted from the SULFO-TAG. Results for the V-PLEX SARS-CoV-2 panel 2 are reported in AU/ml and derived from back fitting the measured signals for samples to a calibration curve generated for each plate. The cut-offs for binding antibodies were provided by the manufacturer and applies for venous serum samples. ACE-2 results are reported as percent inhibition calculated according to: (1 – (average sample ECL signal) / (average ECL signal of diluent only)) x100. Highly positive samples show high percent inhibition whereas negative or low samples show low percent inhibition.

#### In vitro pseudovirus neutralization assay

SARS-CoV-2 spike-pseudotyped lentiviruses were generated by the co-transfection of HEK293T cells with a spike-encoding plasmid (with a 19 amino acid C-terminal truncation), a lentiviral packaging plasmid (Addgene #8455), and a firefly luciferase-encoding transfer plasmid (Addgene #170674) using polyethylenimine (PEI). Culture media was replaced 12-16 h after transfection, and pseudotyped viruses were harvested from the supernatant at 48- and 72-h post transfection, clarified by centrifugation, and stored at -80C until use. Pseudoviruses viruses titrated to generate ~100,000 relative light units (RLUs) were incubated with serial 3-fold dilutions of heat-inactivated serum for 60 min at 37C, and then ~10,000 HEK293T-hACE2 cells were added to each well. Plates were incubated at 37C for 44-48 h and luminescence was then measured using Bright-

Glo (Promega) on a GM-2000 luminometer (Promega) per the manufacturer's instructions. Neutralization was calculated relative to the mean of eight control wells infected in the absence of antibody and fit using a four-parameter logistic curve in Prism v9 (GraphPad Software).

**Definition of effective vaccination, infections before study start and missing data**

Based on the antibody responses observed in this study, a booster vaccine dose was considered effective at day six after administration. To account for viral RNA persistence, only the first SARS-CoV-2-positive specimen was counted per 90-day period. To account for early infections that were missed in the early pandemic phase before PCR-diagnostics was widely available, we assessed antibodies directed against the SARS-CoV-2 viral nucleocapsid in first capillary blood sample donated by a study subject. An individual with N-directed SARS-CoV-2 antibody titer >5000 was considered to have been infected. The immunological data used were complete except for 3 345 capillary samples lacking manually registered sampling dates. These dates were approximated by using laboratory arrival date and subtract three days for postal service delivery to the laboratory (this was the median time between sampling date and arrival date for 3 567 samples that included sampling and laboratory arrival dates).

**Data processing packages**

The R-packages used were readr (version 2.1.1), tidyverse (version 1.3.1), lubridate (version 1.8.0), magrittr (version 2.0.2), xlsx (version 0.6.5), scales (version 1.1.1), survival (version 3.2 – 13), survminer (version 0.4.9), mgcv (version 1.8 – 38), and car (version 3.0 – 13).

**Statistical analyses**

The generalized additive model with cubic splines was based on 404 paired samples from 202 infection-naïve individuals and were taken between day six after vaccine dose 3 up to the administration date of dose 4. Residual analysis showed good performance of the model and four knots produced the best model with regards to fit and flexibility (Supplemental figure 7). We used 1010 observations of AUC in 1010 infection-naïve study subjects from day 20 to day 80 after dose 3 of in the nursing home cohort to approximate the decline in S-directed antibodies. We applied logarithmic regression with log(AUC) as dependent variable and time in days as explanatory variable. We observed a linear decline in scatterplots of log(AUC) and residual analysis verified normality of residual errors and homogeneity of residual variance (Supplemental figure 8).

For the evaluation of SARS-CoV-2-associated mortality, we included study subjects with at least one valid laboratory result from a blood sample taken at day six or later after vaccine dose 3. This allowed us to project antibody levels to day 60 under the assumption of the same antibody half-life for all individuals. We included all PCR-verified first-time infections at day six post dose 3 or later. These restrictions defined SARS-CoV-2 cases and a control population. We randomly sampled two controls per case at the day of positive PCR sampling for each case. Controls were eligible if alive at the case infection date, they had received an effective vaccine dose 3, and were not infected by SARS-CoV-2 the following 30 days. We used the log-rank test to compare cases and controls under the null hypothesis that both populations have identical hazard-functions. In the Cox proportional hazard model to estimate the risk of death within 30 days adjusting for additional risk factors interaction between responder and covid was tested. No individual was censored.

**Supplemental table 1**

| Factor | Nursing Home residents | Individuals <65 years | Covid-19 experienced |
|---|---|---|---|
| **N** | 96 | 53 | 28 |
| Male (%) | 29 (30) | 18 (34) | 11 (39) |
| Female (%) | 67 (70) | 35 (66) | 17 (61) |
| **Age Median (range)** | 85 (71-100) | 50 (21-60) | 80 (27-97) |
| **Days between dose 3 and sampling Median (range)** | 30 (25-35) | 32 (26-56) | 28 (25-35) |
| **Days between dose 2 and 3 Median (IQR)** | 242 (218-259) | 192 (185-204) | (87-242) |

Characteristics of cohorts for sampling at approximately 30 days post dose 3

**Supplemental table 2**

| | Nursing Home residents | Community living >65 | Community living <65 |
|---|---|---|---|
| **N (sample at least one timepoint)** | 16 | 27 | 5 |
| Male (%) | 3 (19) | 15 (54) | 0 (0) |
| Female (%) | 13 (81) | 13 (46) | 5 (100) |
| **Age (median, range)** | 84 (41-93) | 81,5 (76-91) | 57 (51-61) |
| **Days between dose 2 and 3 Median (IQR)** | 219 (194-225) | 199 (190-203) | 218 (199-309) |
| **Days between dose 3 and 4 Median (IQR)** | 143 (143-144) | 123 (120-125) | 268 (268-271) |

Characteristics of individuals that were sampled at dose 4 and up to 30 days post 4.

**Supplemental table 3.**

| Factor | Hazard ratio | Statistic (Log rank test) | p-value |
|---|---|---|---|
| S IgG AUC <479 at day 60 post dose 3 | 3.61 | 3.94 | <0.0001 |
| Primary infection with SARS-CoV-2 | 2.70 | 3.96 | <0.0001 |
| Male gender | 2.19 | 2.36 | 0.0086 |
| One year increase in age | 1.04 | 2.30 | 0.0216 |

Cox proportional-hazards model to investigate factors associated with 30-day mortality in the study population.

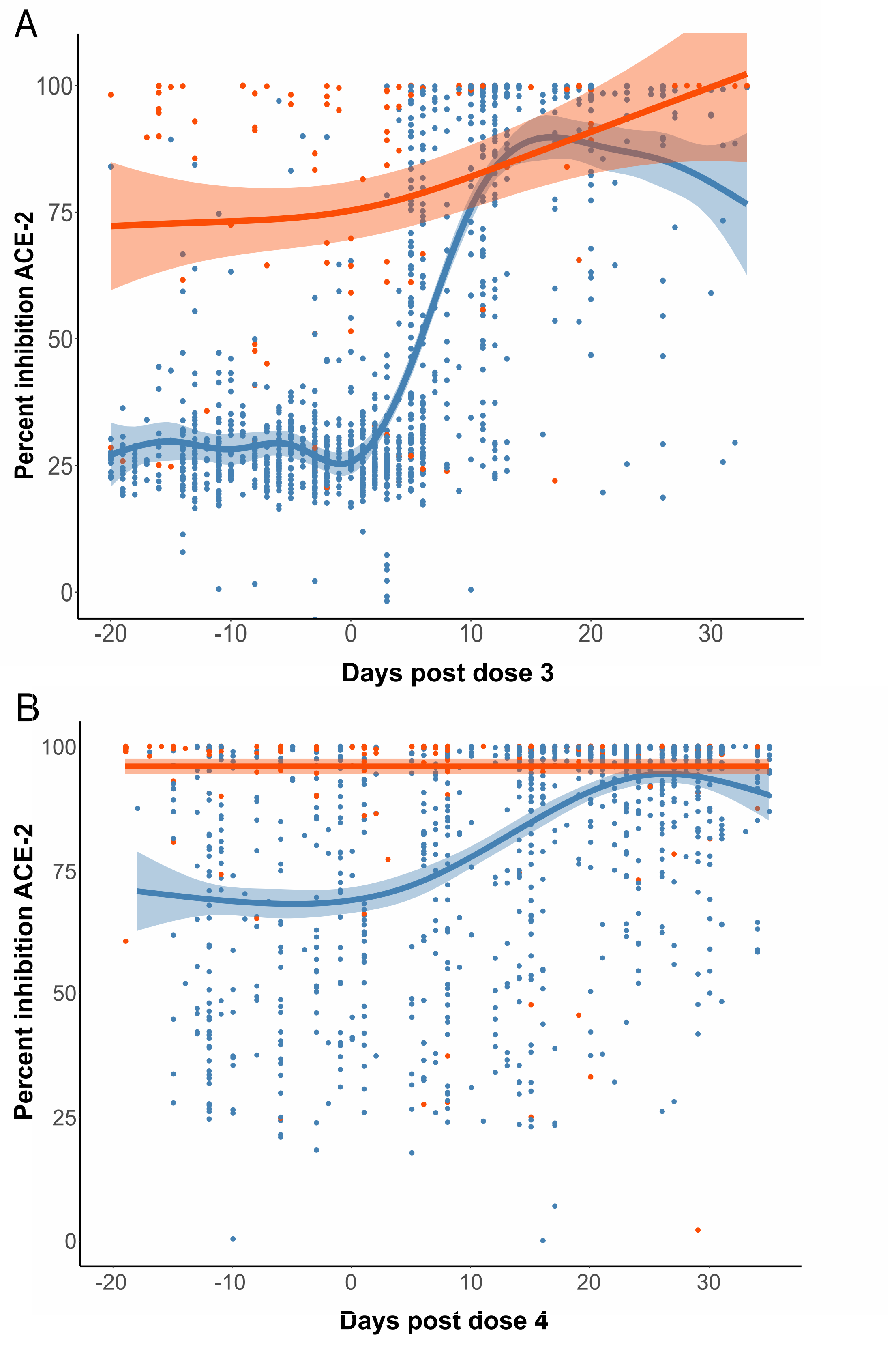


**Supplementary figure 1.** Pseudo neutralization after mRNA vaccine dose 3 (panel A) and dose 4 (panel B). Day 0 indicates the day of vaccination. Dots represent individual observations. Regression lines with 95% intervals are shown. Blue color indicates no previous SARS-CoV-2 infection. Red color indicated that there was a history of previous PCR-verified infection.

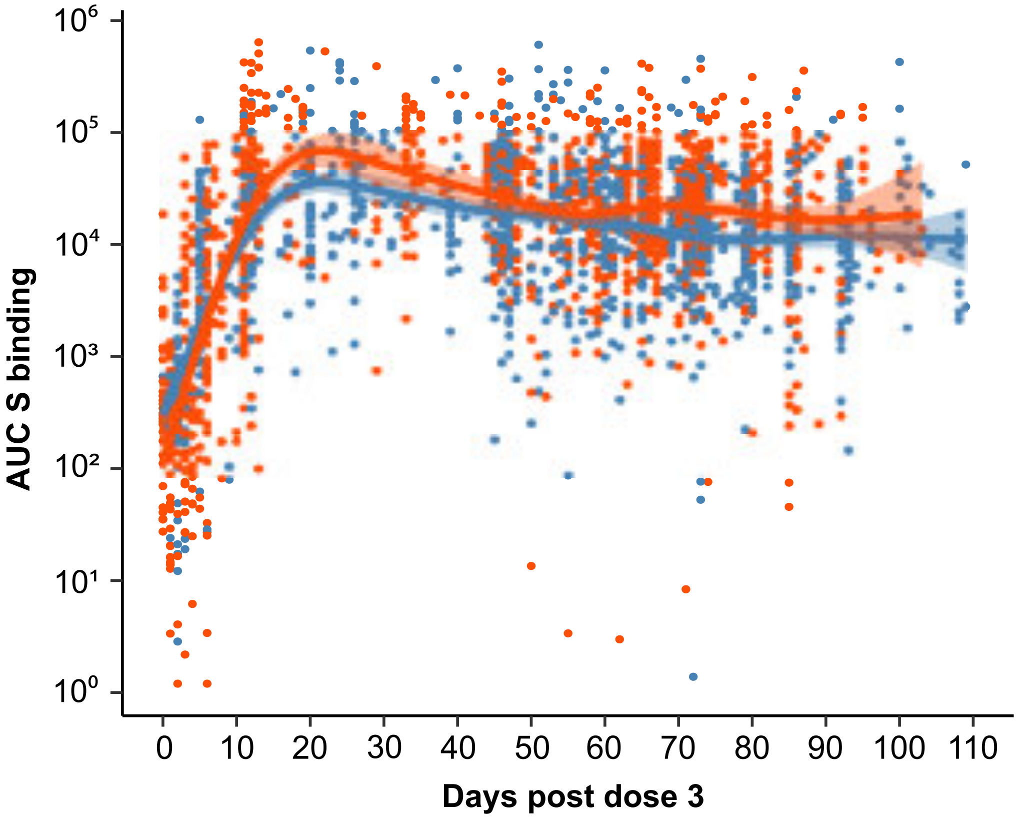


**Supplementary figure 2.** Trends of S-directed IgG in sera by vaccine type. A third vaccination by a half-dose of mRNA-1273 (red) was different from a dose of BNT162b2 (blue). Observations are shown as dots with colored lines representing a cubic spline model with 8 knots to capture trends with 95% confidence intervals. Day 0 indicates the day of vaccination.

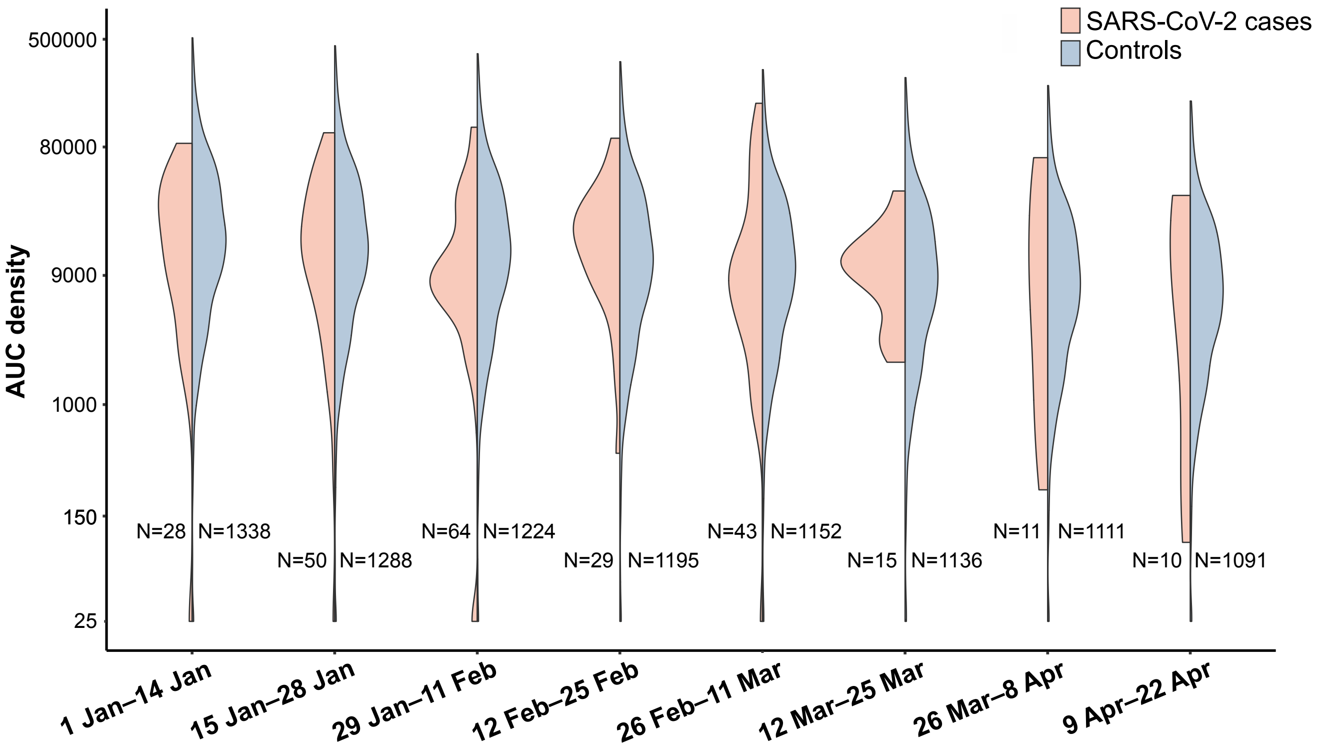


**Supplementary figure 3.** Projected populaton distributon of S-binding IgG among SARS-CoV-2 infected (red) and non-infected (blue) nursing home residents during the Omicron wave. Antibody levels are shown asdensity plots per 14-day period starting 1 January 2021. All included study subjects were previouslyuninfected with SARS-CoV-2 and had received a third mRNA vaccine dose.

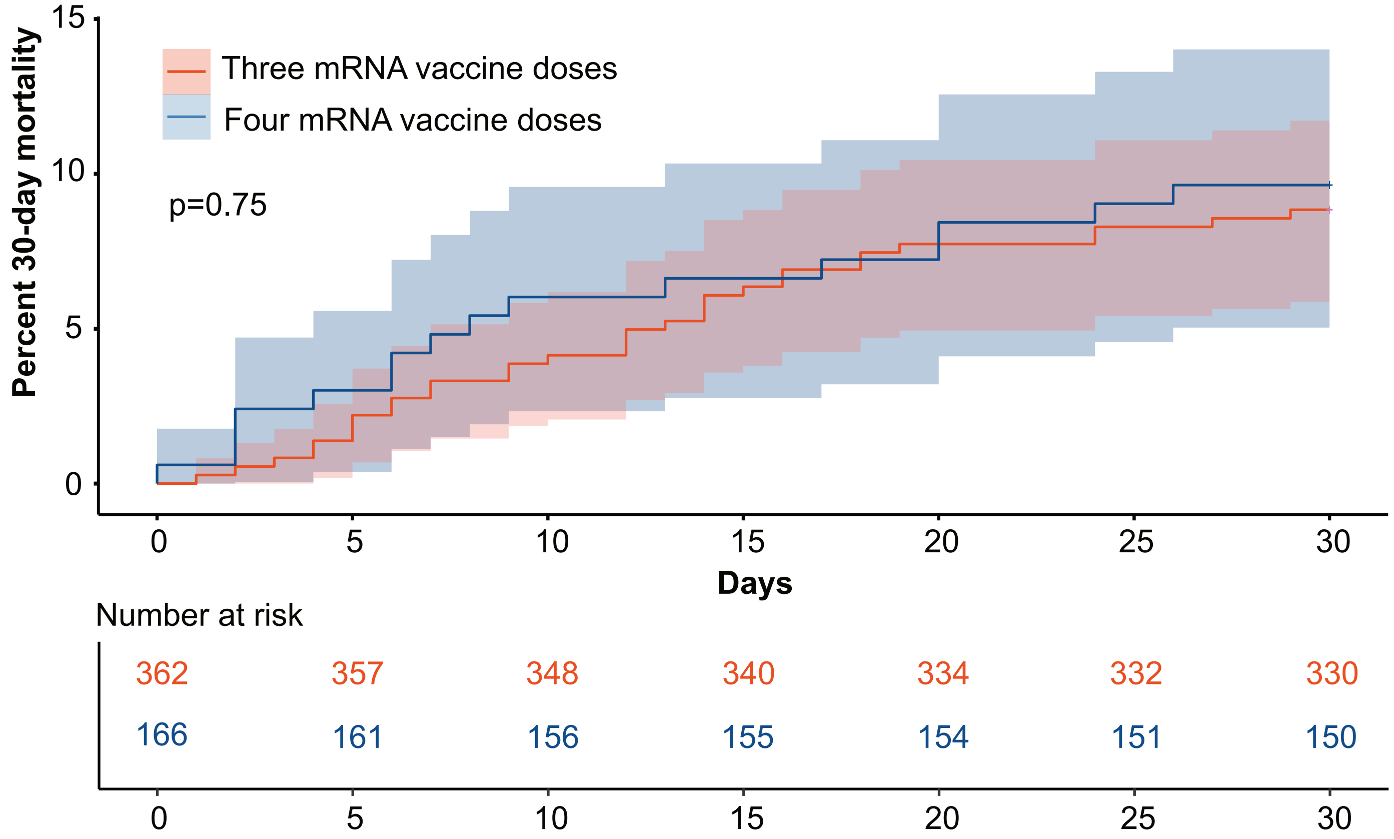


**Supplementary figure 4.** Deaths among SARS-CoV-2-infected study subjects at nursing homes. Cumulative 30-day SARS-CoV-2-associated mortality among 528 previously infection naïve nursing home residents that were vaccinated with three (red) of four (blue) mRNA vaccine doses.

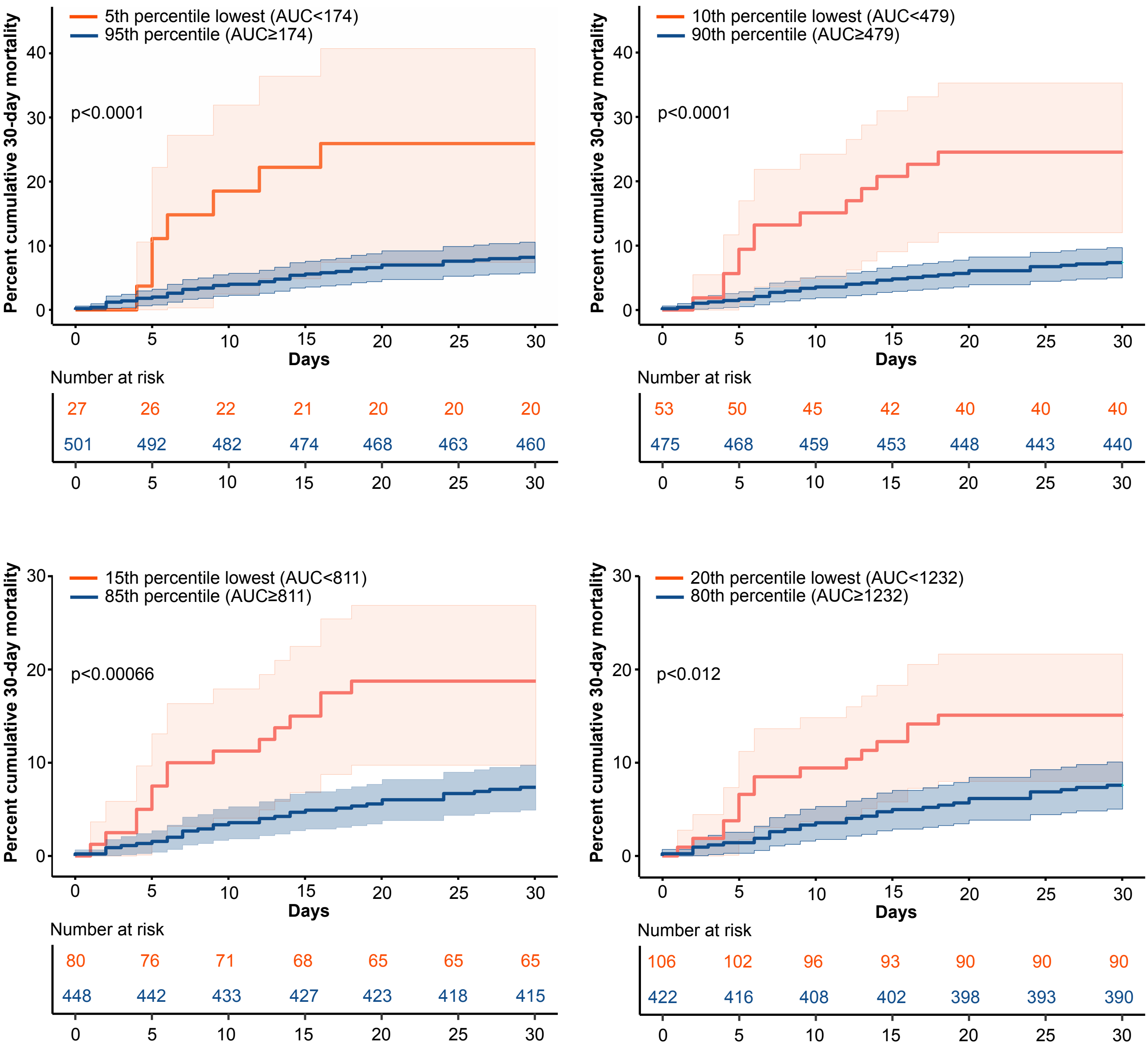


**Supplementary figure 5.** Relationship between S-directed IgG antibody levels at day 60 after vaccine dose 3, and 30-day mortality among 528 SARS-CoV-2-cases. The four panels show the infected population divided by the lowest percentile of antibody levels (5, 10, 15 and 20 percent lowest). Kaplan-Meier curves with 95% confidence intervals for the lowest percentile (red) and the remaining population (blue) are shown. Day 0 is the day of the PCR-verified SARS-CoV-2 infection.

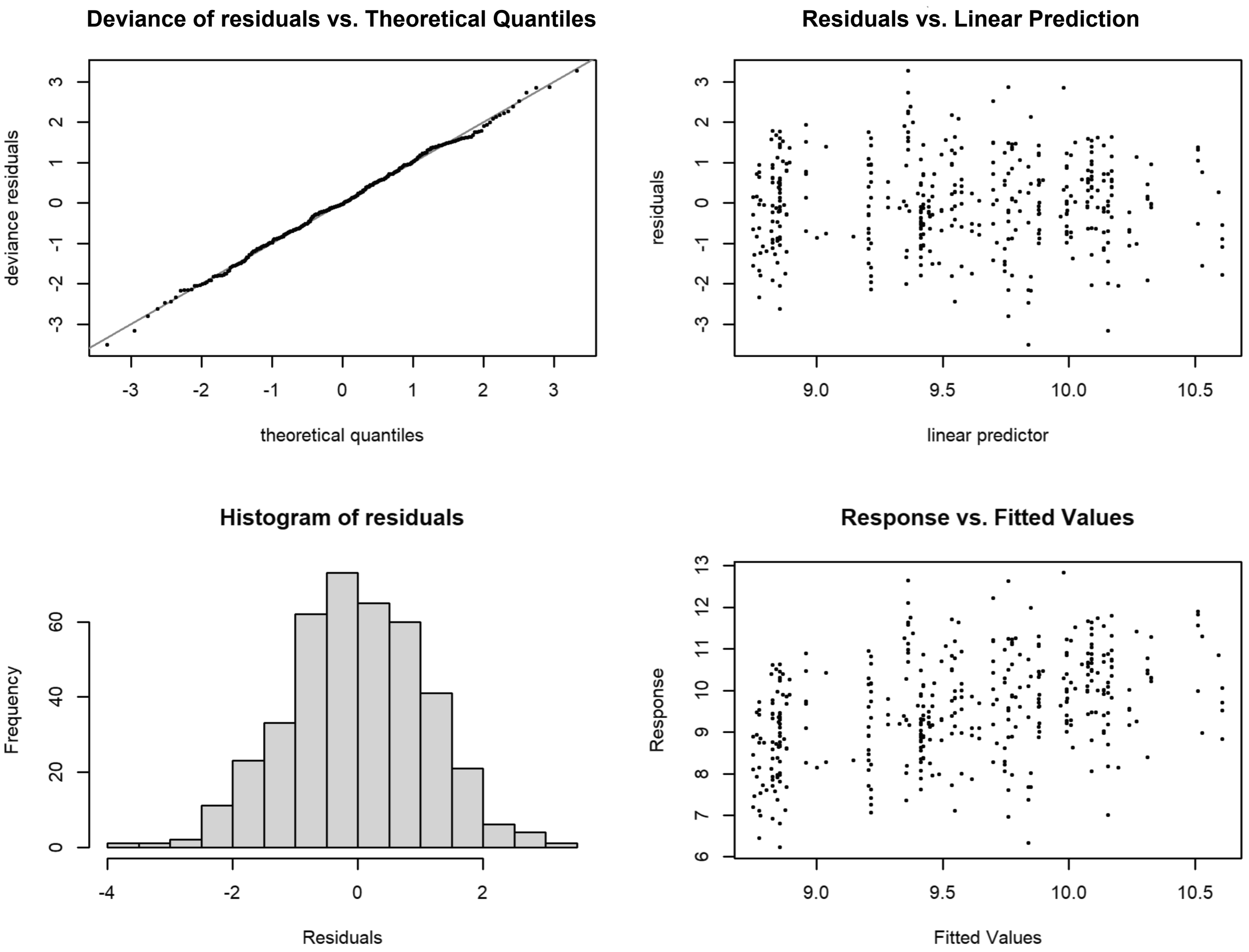


**Supplementary figure 6**. Model tests for the generalized additive model used for estimating differences between vaccine brands. Analyses tested for gender difference, interaction between vaccine brands and the pace of decline in AUC over time by analyzing S-directed antibody AUC-levels after dose 3 in paired samples from 202 study subjects with no SARS-CoV-2-infection between the samples and no history of previous infection.

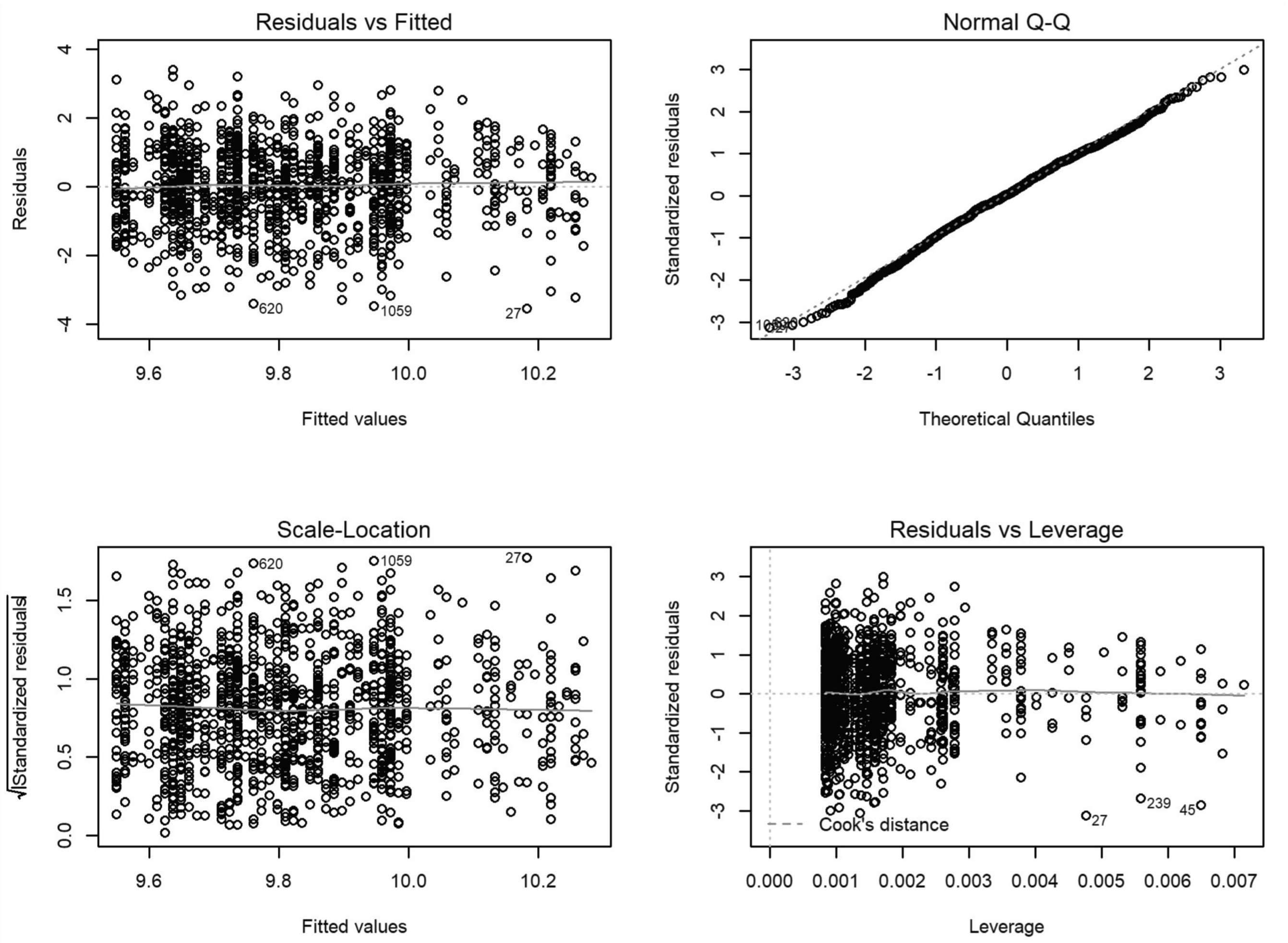


**Supplementary figure 7**. Model tests for a logarithmic regression model used for approximating the decline in AUC-levels. The analyses included 1010 observations of 1010 unique individuals from day 20 to day 80 after vaccine dose 3.
